# The Omega-3 Index is Inversely Associated with the Neutrophil-Lymphocyte Ratio in Adults

**DOI:** 10.1101/2021.10.22.21264656

**Authors:** Michael I. McBurney, Nathan L. Tintle, William S. Harris

## Abstract

The neutrophil-lymphocyte ratio (NLR) is a biomarker of systemic inflammation and measures innate-adaptive immune system balance. The omega-3-index (O3I) measures the amount of EPA+DHA in blood. Both a low O3I and an elevated NLR are associated with increased risk for chronic disease and mortality, including cardiovascular diseases and cancer. Hypothesizing that low O3I may partly contribute to systemic chronic inflammation, we asked if a relationship existed between O3I and NLR in healthy adults (≥18y, n=28,871, 51% female) without inflammation [C-reactive protein (CRP) <3mg/mL)] who underwent a routine clinical assessment. NLR was inversely associated with O3I before (p<0.0001) and after adjusting for age, sex, BMI, and CRP (p<0.0001). Pearson correlations of other variables with NLR were r=0.06 (CRP), r=0.14 (age), and r=0.01(BMI). In this healthy population, an O3I <6.6% was associated with increasing NLR whereas NLR remained relatively constant (low) when O3I >6.6%, suggestive of a quiescent, balanced immune system.

## 1. Introduction

Cells from both the innate and adaptive immune system circulate in blood. Cells of the former (i.e., neutrophils, phagocytes, dendritic cells, macrophages, eosinophils, basophils, mast cells, and natural killer cells) work together to neutralize pathogens, in part by releasing pro-inflammatory mediators that modulate the latter. Neutrophils are the most abundant leukocytes in circulation. They migrate to sites of tissue injury or damage and are responsible for a non-specific inflammatory response. The adaptive system (acquired immunity) responds to specific pathogens and cytokines by producing antibodies and maintains immunological memory via lymphocytes, i.e., T and B cells. The hallmark of the adaptive immune system is clonal expansion of lymphocytes. Inflammation is generally self-limiting, with its resolution being an active rather than passive process [1]. Chronic or unresolved inflammation can damage host tissues and increase risk for the development of a range of non-communicable diseases, e.g. cardiovascular, cancer, chronic respiratory diseases, diabetes, and autoimmune disorders, e.g. rheumatoid arthritis [2–4].

The neutrophil-lymphocyte ratio (NLR) is an emerging biomarker of systemic inflammation [5–8] and innate-adaptive immune system balance [6,9] that is readily accessible from a complete blood count. It has the benefit of not being influenced by physiological conditions such as dehydration and exercise [6]. NLR has been used to monitor astronaut immune function during long-duration missions [10]. Elevated NLR is also a biomarker of systemic inflammation [5,11–14], cardiovascular disease and events [6,15–18], the severity of covid-19 [19–22], mood disorders [23], cognitive impairment [24], cancer [6,7,25], dry eye [26], periodontitis [27], rheumatoid arthritis [28], cancer [12,25,29], diabetic kidney disease [30] and total mortality [6,9,14,17,31,32].

Nutrient deficiencies or inadequacies can impair immune system function and weaken the immune response [3,33–36]. The proportion of EPA and DHA in immune cell membranes can modulate inflammation via the synthesis of prostaglandins, leukotrienes, lipoxins and specialized pro-resolving lipid mediators (SPMs, e.g., resolvins, protectins, and maresins) [1,37]. The EPA and DHA-derived metabolites are less inflammatory than those derived from the omega-6 precursor, arachidonic acid [1,33,37–42]. SPMs derived from EPA and DHA reduce neutrophil infiltration and production of reactive oxygen species, regulate the cytokine-chemokine axis, and temper the inflammatory response without immunosuppression [37,43– 45]. The omega-3 index (O3I) [46] is a stable biomarker of long term EPA and DHA intake [47,48]. A low O3I [49] and an elevated NLR [14,18,50] are both associated with risk of cardiovascular events and mortality (**Supplemental Table 1**). Hypothesizing that low blood EPA+DHA levels may partly contribute to systemic chronic inflammation, we asked if there was a relationship between the O3I and the NLR in healthy individuals without inflammation.

## 2. Patients and Methods

This is a cross-sectional analysis of data from blood samples submitted for testing to Health Diagnostic Laboratory, Inc (HDL, Inc., Richmond VA; now defunct) as part of routine clinical assessment between 2011-2012. Subjects were adults (≥18y) with data on O3I, NLR, body mass index (BMI), age, sex, and high-sensitivity C-reactive protein (CRP) (n=44,925). Individuals with extreme O3I values (i.e., highest and lowest 0.5%) were excluded. Because CRP > 3mg/L is indicative of inflammation arising from infection, trauma or chronic disease [51–54], individuals with CRP >3 mg/L were also excluded. Thus, the final study population consisted of 28,871 individuals (**Figure 1**).

**Figure 1.**
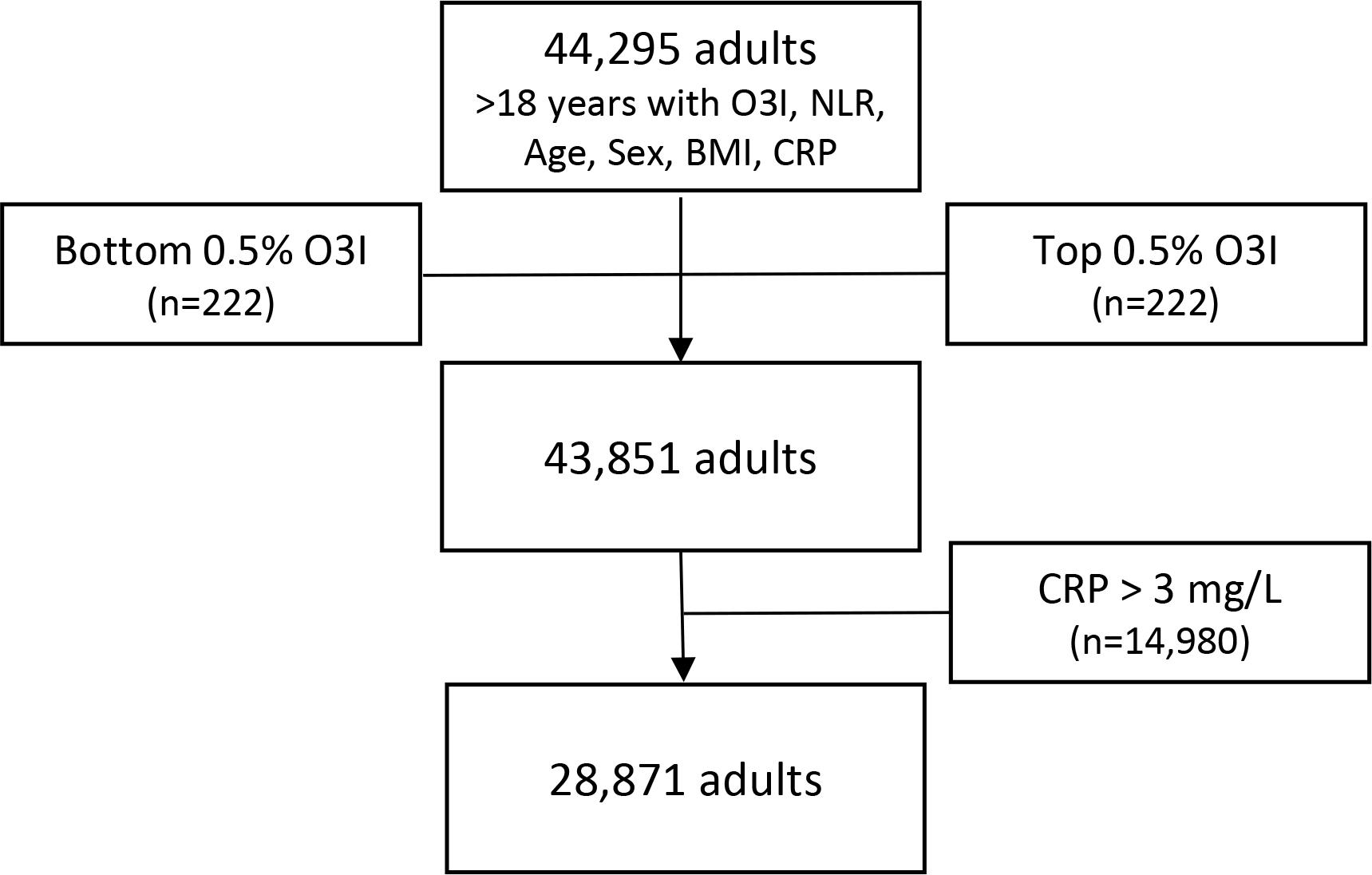
Analytical sample flow. NLR, neutrophil-lymphocyte ratio; O3I, omega-3 index; high-sensitivity C-reactive protein; BMI, body mass index.

### 2.1 Laboratory Methods

Blood samples were drawn after an overnight fast and shipped with cold packs to HDL, Inc. for testing. Samples were prepared at each clinical site according to standardized instructions as previously described [55]. Absolute concentrations of blood cells (neutrophils, lymphocytes, NLR) were determined with a Beckman-Coulter DxH 800 analyzer (Brea, CA, USA) and biomarker data (CRP, O3I) were extracted without any linked patient identifiers except age, sex, and BMI. For fatty acid analysis, RBCs were separated from plasma by centrifugation and analyzed using gas chromatography as previously described [56]. The University of South Dakota Institutional Review Board reviewed and approved the use of such de-identified clinical data for research purposes (IRB-21-147).

### 2.2 Statistical methods

Sample characteristics were summarized using standard statistical methods (e.g., means, SDs, correlations) with t-tests and adjusted linear models used to compare characteristics of male and female participants above and below O3I values that were ultimately identified as primary cut points in the NLR-O3I curves (see below). Splines were fit using 3rd degree polynomials with knots at each decile in R (version 3.6.2; splines package). Unadjusted models used a linear model to predict NLR values by splines of O3I. Adjusted models accounted for sex, age, BMI and CRP values in the linear models accounting for potential non-linear relationships using splines. To identify significant changes in the NLR-O3I relationship, we used a “sliding O3I window” approach. The width of each window was three O3I percentage points (e.g., 4% to 7%). By moving the window up by 0.1% increments and repeatedly testing for significant differences between the mean NLR in the lower vs. the upper half of the window, we sought to discover O3I cut points where the NLR-O3I relationship appeared to flatten. These would be O3I values above which the “effect” of an increase in O3I on NLR had little impact. We began with a window midpoint of O3I =2.6%, and we used adjusted linear models in R to test for upper vs lower half differences. We identified the first window which did not have a statistically significant difference in upper and lower mean NLR values. The midpoint of this window was chosen as the O3I cut point to be used in further analysis.

Statistical interactions with O3I values above and below the subsequently identified inflection points were tested by placing the interaction term in separate models predicting O3I, NLR, age, sex, BMI, and CRP. Pearson correlations were used to assess strength and direction of linear association between covariates and NLR. Statistical significance was set to 0.05 for all analyses and 95% confidence bands are provided where appropriate.

## 3. Results

The final dataset consisted of 28,871 healthy adults (**Figure 1**). The average age was 55.1±15.1 years, the average BMI was 27.6±5.4 kg/m^2^, and there were slightly more females (51%) than males. Females were significantly younger and had slightly but significantly higher O3I and CRP levels and lower NLR, neutrophil count, and BMI than males (**Table 1**). When the model was also adjusted for age, BMI and CRP, small but significant sex differences remained with the exception of BMI (**Table 1**).

**Table 1.**
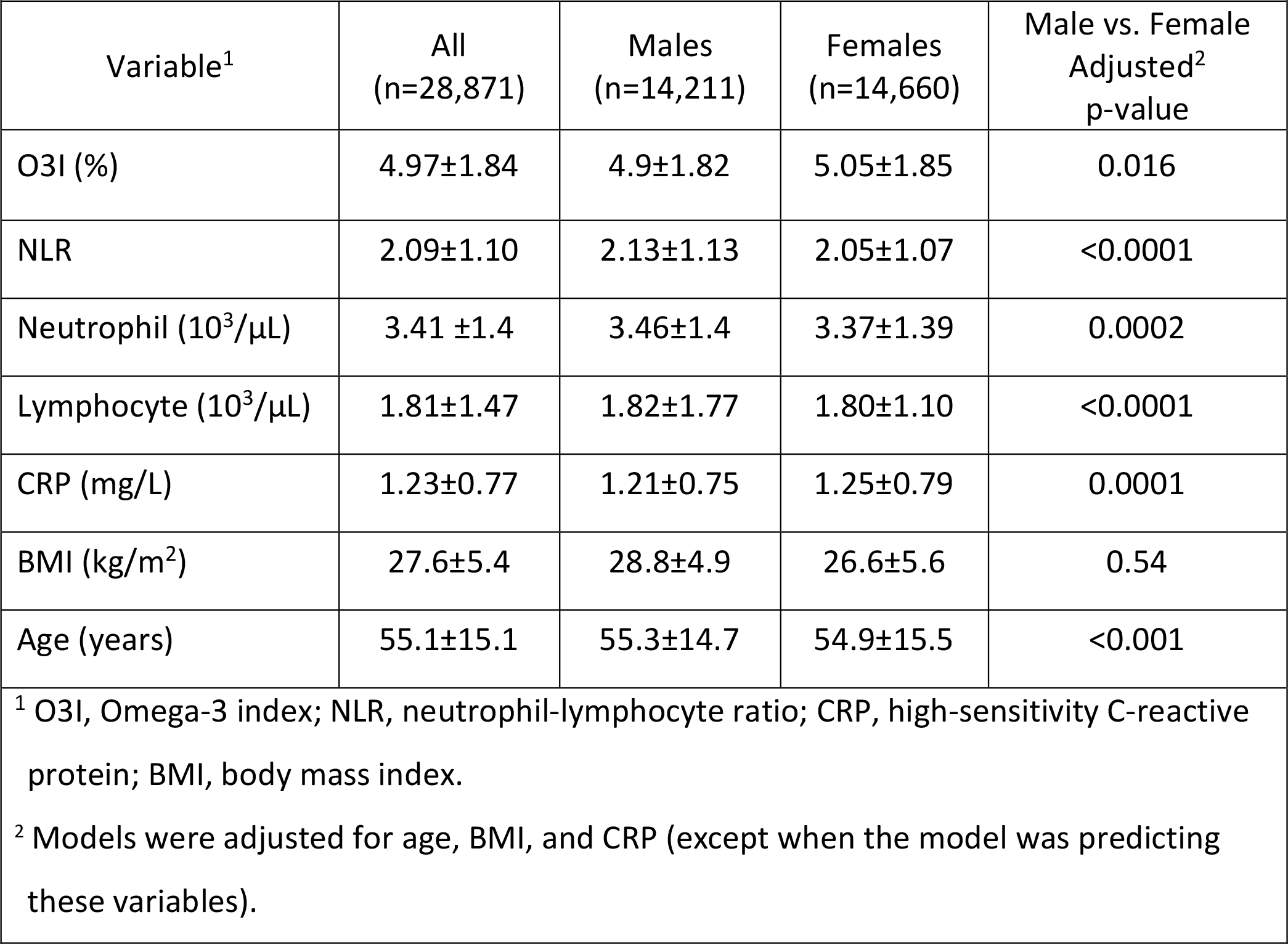
Characteristics of the study population. Mean±SD

NLR was significantly (p<0.0001) and inversely associated with O3I (**Figure 2A**). Pearson correlations (r) of other variables with NLR were r=0.06 (CRP), r=0.14 (age), r=0.01 (BMI), r=0.65 (neutrophil count), and r=-0.24 (lymphocyte count). Adjustment for age, sex, BMI, and CRP strengthened the relationship (**Figure 2B**). The O3I value where the relationship between O3I and NLR began to flatten (i.e., the cut point determined by the sliding window analysis described in Methods) was 6.6% although NLR continued to decline until O3I >8.5% (**Figure 2B**). Below 6.6%, the curve was clearly steeper than it was above 6.6%. The O3I cut point of 6.6% was explored in more detail. An O3I <6.6% was associated with significantly higher NLR, neutrophil number, lymphocyte number, CRP, BMI, and lower age (versus O3I ≥6.6%) (**Table 2**). These O3I-related differences persisted, except for lymphocyte number, when the model was adjusted for age, sex, BMI and CRP (**Table 2**).

**Table 2.** Stratification by omega-3 index (O3I) classification (n=28,871). Mean±SD

|  |  |  |  |
| --- | --- | --- | --- |
| Variable<sup>1</sup> | O3I <6.6%<br>(n=23,510) | O3I ≥6.6%<br>(n=5,361) | Adjusted<sup>2</sup><br>p-value |
| O3I (%) | 4.28±1.11 | 8.03±1.16 | <0.0001 |
| NLR | 2.12±1.11 | 1.96±1.07 | <0.0001 |
| Neutrophil (10<sup>3</sup>/μL) | 3.48±1.40 | 3.11±1.35 | <0.0001 |
| Lymphocyte (10<sup>3</sup>/μL) | 1.82±1.56 | 1.76±0.95 | 0.591 |
| CRP (mg/L) | 1.27±0.78 | 1.07±0.73 | <0.0001 |
| BMI (kg/m<sup>2</sup>) | 28.0±5.4 | 26.3±4.9 | <0.0001 |
| Age (years) | 54.1±15.4 | 59.4±13.1 | <0.0001 |
| <sup>1</sup> O3I, Omega-3 index; NLR, neutrophil-lymphocyte ratio; CRP, high-sensitivity C-reactive protein; BMI, body mass index. |  |  |  |
| <sup>2</sup> Models were adjusted for sex, age, BMI, and CRP (except when the model was predicting these variables). |  |  |  |

**Figure 2.**
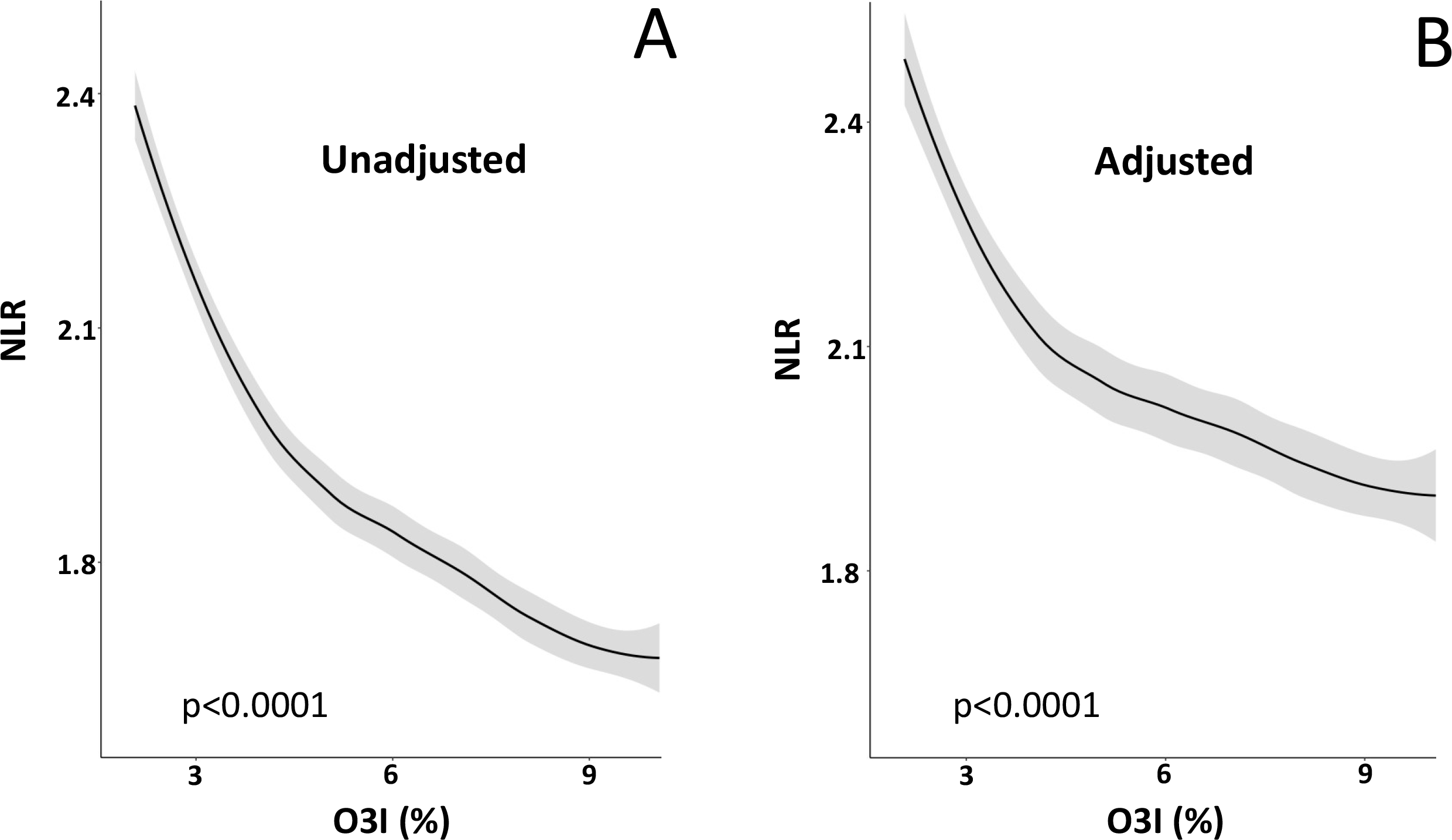
The unadjusted (A) and adjusted for sex, age, BMI, and CRP (B) relationship between the neutrophil-lymphocyte ration (NLR) and the omega-3 index (O3I) in 28,871 healthy adults. (Means and 95% confidence bands).

When adults with inflammation (CRP >3mg/L) were included (n=43,851), NLR was significantly and inversely related with O3I before (p<0.0001) and after adjusting for age, sex, BMI and CRP (p<0.0001) (**Supplemental Figure 1**). As noted above, we chose to restrict our analysis to a healthy population (i.e., without evident inflammation) to establish EPA+DHA standards that could be used to develop DRIs.

## 4. Discussion and Conclusions

We report a significant inverse relationship between O3I and NLR. By excluding individuals with CRP >3 mg/L, we could examine the O3I-NLR relationship in individuals without any evidence of tissue injury, infection, or systemic acute inflammation [57,58], i.e. a cohort suitable for deriving O3I standards on which Dietary Reference Intakes (DRIs) could be developed for planning and assessing EPA+DHA intakes of healthy people. NLR values measured in this cohort (**Table 1**) were similar to those measured in healthy adults globally [8,24,59–63]. NLR is known to increase with age [9,32,60], obesity [13,60], CRP [64,65], and to sometimes differ by sex [32,59]. These same relationships were mostly observed here as well (data not reported), suggesting that the findings from this clinical laboratory cohort are generally representative.

Inflammation is a normal physiological response to infection and injury, but unrestrained inflammation can cause tissue damage leading to disease. As noted earlier, EPA and DHA serve as precursors for the synthesis of prostaglandins, leukotrienes, lipoxins and SPMs that can mediate immune responses [1,29,33,37,39–41]. SPMs derived from EPA and DHA protect tissues by limiting acute inflammatory responses and helping achieve homeostasis without immunosuppression [37,44,45]. In a randomized, double-blind, crossover trial, supplementation (10wk) with high doses of EPA or DHA (∼8g/d) in individuals with abdominal obesity and low-grade inflammation (baseline CRP 3.3±2.4 mg/L) separated by a 9-wk washout significantly increased blood EPA and DHA concentrations (vs corn oil control) [66]. DHA, but not EPA, supplementation decreased CRP concentrations (vs control) and the authors concluded EPA and DHA had more consistent effects on anti-inflammatory gene expression than pro-inflammatory gene expression [66]. Using pairwise and network meta-analyses of randomized controlled trials, DHA and EPA seem to have similar effects on CRP, IL-6, TNF-α, and adiponectin [67]. Although EPA and DHA trigger anti-inflammatory responses in vitro [68,69], the effects of long-chain omega-3 fatty acids on plasma CRP concentrations are mixed [70–73]. A threshold blood EPA+DHA concentration may be required to fuel sufficient SPM biosynthesis to achieve meaningful resolution of inflammation [37].

Omega-3 supplementation increases the EPA+DHA content of neutrophil membranes in vivo and inhibits the 5-lipoxygenase pathway and leukotriene B4-mediated functions of neutrophils *in vitro* [38]. Omega-3 supplementation significantly increased plasma EPA+DHA levels (50-100%) and decreased concentrations of IL-6 (10-12% vs 36% increase in placebo group) and TNFα (0.2 to −2.3% vs 12% increase in placebo group) in healthy, middle-aged and older individuals [74]. Meta-analyses have confirmed this effect [72]. In a study of 8,237 participants living in Japan without a history of cardiovascular disease, eating fish >4 days per week was associated with significant lower NLR [75]. High omega-3 dietary intake was associated with a trend towards lower CRP (p=0.09) and NLR (p=0.17) in men with acute coronary syndrome [76]. A systematic review and meta-analysis of 18 randomized controlled trials found marine-derived omega-3 fatty acids lowered pro-inflammatory eicosanoid concentrations, e.g., in neutrophil leukotriene B4 [77]. In the long-term, chronically lower levels of inflammation may have health benefits. For example, a pooled analysis of data from 17 prospective cohort studies involving 15,720 deaths among 43,466 individuals over a median of 16 years of follow-up found blood EPA+DHA levels were inversely associated with risk for death from all causes, CVD, cancer and other causes [49].

Despite the strength of the NLR-O3I relationship, the most obvious limitation of this study is that confounding metabolic events, dietary behaviors, smoking habits and/or physical activity levels associated with O3I may also affect innate-adaptive immune system balance. However, a nationally representative study of 9,427 individuals living in the US did not find significant differences in NLR with respect to sex, education, insurance status, or drinking habits [59]. Moreover, this cross-sectional population is similar to 2/3 of the US population who have CRP <3mg/L [78] and the ∼95% of adults living in the USA with circulating O3I percentages below 5% [79,80].

In this cross-sectional study of healthy individuals, 81% of participants had O3I <6.6%. Below this threshold, an increase in NLR, reflecting an increasingly imbalanced innate-adaptive immune system, was observed. NLR continued to decrease with increasing O3I until O3I >8.5%, similar to the target proposed for cardiovascular health, i.e. O3I >8% [46,81]. Clearly additional evidence, including intervention studies, is needed to determine whether the NLR-O3I relationship is causal or coincidental.

In conclusion, red blood cell EPA+DHA levels are significantly and inversely associated with NLR, a biomarker of inflammation and innate-adaptive immune system balance. Based on our observations in this healthy cohort, an O3I >6.6%, and possibly as high as 8.5%, is associated with lower NLR values that are indicative of a quiescent, balanced innate-adaptive immune system.

## Data Availability

Pending application and approval, data described in the manuscript, code book, and analytic code will be made available upon request to the Fatty Acid Research Institute.

https://www.faresinst.org/

## Abbreviations

BMI: body mass index
CRP: high-sensitivity C-reactive protein
DHA: docosahexaenoic acid
EPA: eicosapentaenoic acid
NLR: neutrophil-lymphocyte ratio
O3I: omega-3 index
SPM: specialized pro-resolving lipid mediators.

## Acknowledgements

**T**he authors wish to thank Steven Varvel, PhD for his help in acquiring and collating the dataset used in this study.

## CReditT authorship contribution statement

**M**.**I. McBurney**: Conceptualization & analytical design. Writing -original draft, review & editing. **W**.**S. Harris**: Conceptualization & analytical design. Funding acquisition. Writing- review & editing. **N**.**L. Tintle**: Formal analysis. Writing-review & editing.

