## Supplemental Table 1 and Figure 1. for "The Omega-3 Index is Inversely Associated with the Neutrophil-Lymphocyte Ratio in Adults"

[1]Fatty Acid Research Institute, Sioux Falls, SD 57106, USA (MIM, NLT, WSH); [2]Department of Human Health and Nutritional Sciences, University of Guelph, Guelph, ON N1G 2W1, Canada (MIM), [3]Division of Biochemical and Molecular Biology, Friedman School of Nutrition Science and Policy, Tufts University, Boston, MA 02111, USA (MIM); [4]Department of Population Health Nursing Science, College of Nursing, University of Illinois – Chicago, Chicago, IL 60612, USA(NLT); [5]Sanford School of Medicine, University of South Dakota, Sioux Falls, SD 57105, USA (WSH).

| Supplemental Table 1. Associations of a low neutrophil-lymphocyte ratio (NLR) and a low Omega-3 Index (O3I) with adverse clinical events. | | | | |
|---|---|---|---|---|
| Condition or Outcome | Lower NLR | | Higher O3I | |
| | Setting | Finding | Setting | Finding |
| Dry Eye Disease | Case Control | Less likely to be a case [1] | Meta-analysis RCTs | Improves [2] |
| Inflammatory markers | 389 in smoking cessation study | Lower biomarker levels [3] | Framingham | Lower levels [4] |
| CV Events | Meta-analysis of lipid RCTs | Predicted lower event rates regardless of treatment [5] | Meta-analysis of om3 RCTs | Reduced Risk [6] |
| Cancer mortality | NHANES (within 1 year of death) | Lower odds of dying [7] | FORCE meta-analysis | Reduced Risk [8] |
| All-cause mortality | Jackson Heart Study | Lower risk for death [9] | FORCE meta-analysis | Reduced Risk [8] |
| CHD mortality | NHANES | Improves on FRS prediction [10] | FORCE meta-analysis | Reduced Risk [8] |
| HF morbidity and mortality | 1212 patients with acute HF | Lower risk for death [11] | GISSI-HF trial<br>CHS observational study | Reduced mortality [12, 13] and HF incidence [14] |
| Depression | Meta-analysis cases vs controls | Lower risk [15] | Meta-analysis cases vs controls | Reduced risk [16] |
| COVID-19 | Chinese patients | Better outcomes [17] | 100 US patients<br>79 Chilean patients | Reduced risk for death [18, 19] |
| Ischemic stroke | Retrospective in 25K Koreans | Lower risk [20] | 3 US prospective cohorts and Chinese biobank | Lower risk [21, 22] |

Figure 1. The unadjusted (A) and adjusted for sex, age, BMI, and CRP (B) relationship between the neutrophil-lymphocyte ration (NLR) and the omega-3 index (O3I) in 43,851 adults. (Means and 95% confidence bands).

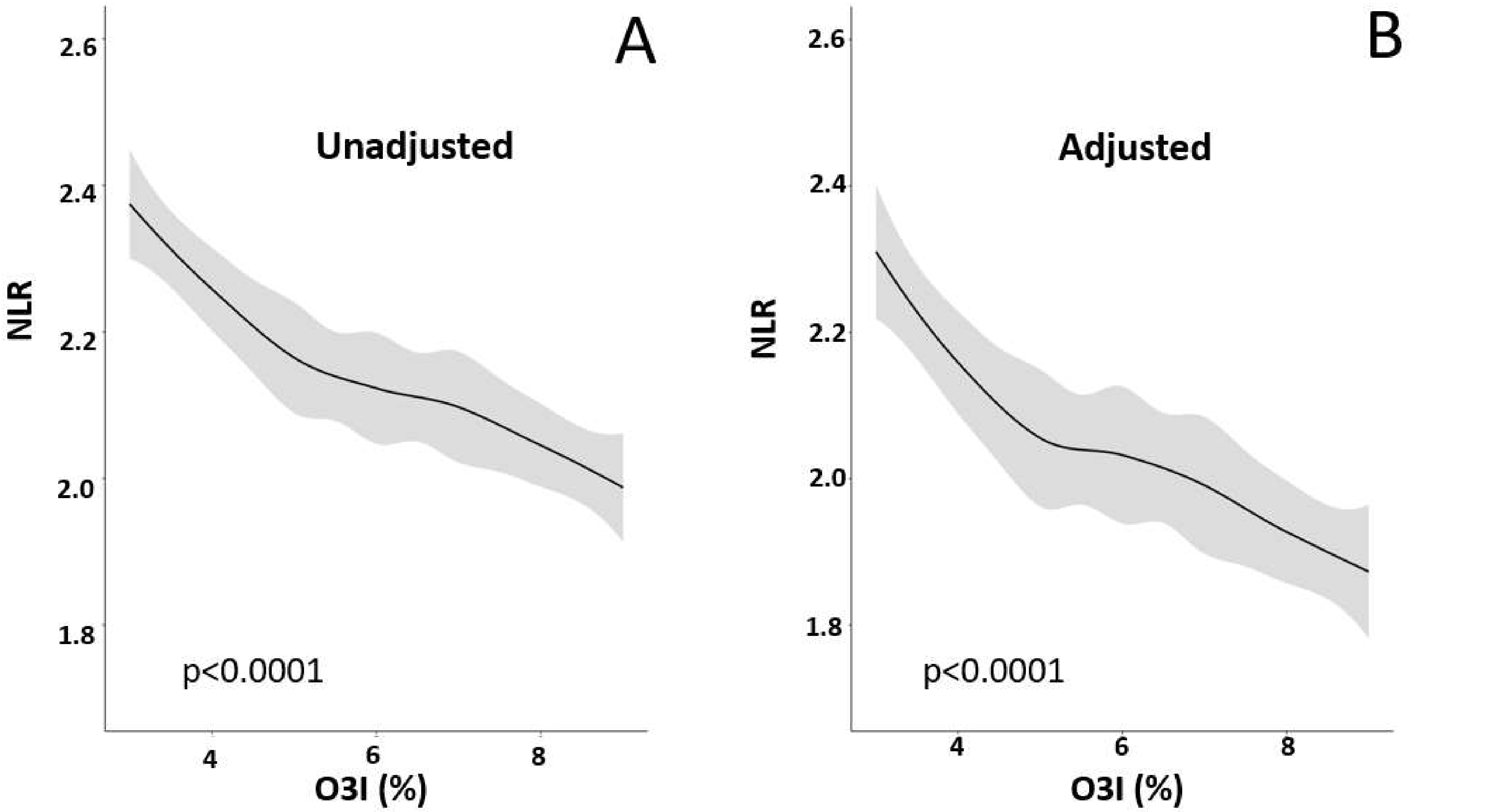
